# DBS for substance use disorders? An exploratory qualitative study of perspectives of people currently in treatment

**DOI:** 10.1101/2022.04.21.22273594

**Authors:** Erika Versalovic, Eran Klein, Sara Goering, Quyen Ngo, Kate Gliske, Marion Boulicault, Laura Specker Sullivan, Mark J Thomas, Alik S Widge

## Abstract

**Objective:** While previous studies have discussed the promise of deep brain stimulation (DBS) as a possible treatment for substance use disorders (SUDs) and collected researcher perspectives on possible ethical issues surrounding it, none have consulted those with SUDs themselves. We addressed this gap by interviewing those with SUDs.

**Methods:** Participants viewed a short video introducing DBS, followed by a 1.5 hour semi-structured interview on their experiences with SUDs and their perspective on DBS as a possible treatment option. Interviews were analyzed by multiple coders who iteratively identified salient themes.

**Results:** We interviewed 20 people in 12-step based, inpatient treatment programs (10 [50%] white/Caucasian, 7 Black/African American [35%], 2 Asian [10%], 1 Hispanic/Latino [5%], and 1 [5%] Alaska Native/American Indian; 11 [45%] women). Interviewees described a variety of barriers they currently faced through the course of their disease that mirrored barriers often associated with DBS (stigma, invasiveness, maintenance burdens, privacy risks) and thus made them more open to the possibility of DBS as a future treatment option.

**Conclusions:** Individuals with SUDs gave relatively less weight to surgical risks and clinical burdens associated with DBS than previous surveys of provider attitudes anticipated. These differences derived largely from their experiences living with an often-fatal disease and encountering limitations of current treatment options. These findings support the study of DBS as a treatment option for SUDs, with extensive input from people with SUDs and advocates.

## INTRODUCTION

Deep brain stimulation (DBS) is a rapidly expanding treatment option for a wide range of conditions, including Parkinson’s disease, obsessive-compulsive disorder, and epilepsy. DBS is currently being explored as a possible treatment for substance use disorders (SUDs) (Wang et al. 2018). Case reports of experimental DBS in SUDs describe notable improvements (Qu et al. 2019; Gonçalves-Ferreira et al. 2016; Muller et al. 2013). Despite promising results from these small samples, enthusiasm has been tempered by concerns from researchers and bioethicists that it may be difficult to study DBS for SUD without coercion (Pisapia et al. 2017). SUDs are socially stigmatized and often criminalized, increasing the risk of people being pressured to pursue treatments to avoid prosecution. It is also unclear whether individuals would accept a physically invasive neurosurgical intervention for SUDs. Past research has argued that SUDs are fundamentally less disabling or dangerous than other DBS indications (Ali et al. 2016; Lee et al. 2016).

That cautious position may not adequately reflect the widespread and severe nature of SUDs, and the limitations of current treatment (Mojtabai et al. 2019, Substance Abuse and Mental Health 2020). While inpatient treatment can help people with SUDs achieve initial abstinence, the rate of relapse is high (Andersson et al. 2019; Gil-Rivas et al. 2009). Agonist therapies are primarily indicated for opioid use disorders, and are often difficult to access. Many of these limitations reflect social stigma and under-investment, and no technology can substitute for the policy solutions needed to reverse structural inequalities. That said, even when patients can access treatment, a substantial fraction cannot recover through existing options alone. For those patients, DBS may be more attractive to people living with SUDs than researchers imagine. Indeed, research target population preferences may differ significantly from researcher or clinician perspectives (e.g. Anderson 2004). Consultation with potential or actual device users can reveal novel considerations about neurotechnology development that can help guide future research (Goering & Klein 2020; Collinger et al. 2013).

Here, we report a qualitative study that explored the values, interests and concerns of people with SUDs in relation to the prospect of DBS as a treatment option. We interviewed people in treatment centers in the early abstinence phase of their SUDs, asking about participants’ experiences with their disorder and treatment, and their perspectives on DBS across five themes: personal agency, social dynamics, stigma, privacy, and interactions with the healthcare system. These aspects reveal how some common concerns surrounding DBS (e.g., physical invasiveness and maintenance burdens) compare to the burdens already experienced through the course of SUDs and their treatment.

## METHODS

We conducted 20 semi-structured interviews with people in residential treatment for SUDs. Interview guides were developed collaboratively through a series of discussions among all authors, building on previous work related to neurotechnologies (Versalovic et al. 2020, Goering et al. 2017, Klein et al. 2016) and with attention to the circumstances of people in treatment for SUDs. Purposive sampling methods were used for representation of a range of substances, and of racial groups often excluded from medical device research (Fox-Rawlings et al. 2018). Participants were recruited through the [anonymized for review] treatment system. The study was reviewed by the [anonymized for review] (STUDY00009975) and [anonymized for review]. Procedures were followed in accordance with both institutional review boards and with the Helsinki Declaration (as revised in 2004) (WMA Declaration of Helsinki).

Participants were initially asked about their experiences with addiction and treatment. They then watched a five-minute video produced by the study team introducing deep brain stimulation (DBS) (see supplemental materials). The interview guide (see supplemental materials) was structured around ethical and social concerns that have arisen in the application of DBS to mental health disorders to explore their saliency within the SUD population. These themes included agency (ex: how could you imagine a DBS enhancing or undermining a user’s sense of agency?), social relationships (ex: would you involve loved ones in the process of getting a DBS?), stigma (ex: how could possible stigma of a neural device interact with stigma surrounding SUDs?), privacy (ex: who should have access to neural data?), and interactions with the healthcare system (ex: what kind of support is needed for follow-up appointments?). At the end of the interview, participants were asked if DBS would be something they might consider if it became available, who they identified as the most appropriate target users (if any), the value of gathering target-user perspectives, and whether they wished to amend any of their responses.

Interviews were conducted and recorded through HIPAA-compliant Zoom by EK and EV and lasted 1.5 hours on average. Demographic surveys were administered online post-interview. Participants were compensated $25 through a gift card. Interviews were transcribed using an online service. SG, EK, and EV read the interviews and conducted thematic content analysis. The first 12 transcripts were each independently, inductively coded on atlas.ti, followed by discussions to reconcile code differences to arrive at the final coding scheme. The final 8 were coded by EV. To ensure sensitivity to the lived experience of SUDs and the treatment process, we used a team-based approach (Giesen and Roeser 2020) with monthly meetings of the full authorial team, including our SUDs subject matter experts, to check in, discuss any difficulties, and make decisions about the research process (e.g., timeline, determining coding schemes, broadening recruitment, etc.). Data were collected from September 2020 to May 2021 and analyzed from May 2021 to December 2021. Methods reported here are in line with the COREQ (Tong et al. 2007) and RATS (Clark 2003) checklists (Neale & West 2015).

## RESULTS

### Demographics

Participant demographics and specifics regarding primary substance, prior treatment, co-occurring disorders, and family history are presented in Table 1. There was nearly equal representation of female and male participants (11:9 respectively). Ten of the 20 participants were white, with seven identifying as Black/African American, two as Asian, one as American Indian/Alaska Native, and one as Hispanic/Latino. There was a spread across ages 25-64 and in education level from some high school to completion of a graduate degree. Participants had backgrounds in a wide range of substances: alcohol being the most prominent (90%), with just under a third identifying marajuana (30%) and with two to three of participants identifying each of the categories of opioids, cocaine, and methamphetamines. 17 of the 20 participants had a family history of SUDs.

**Table 1:**
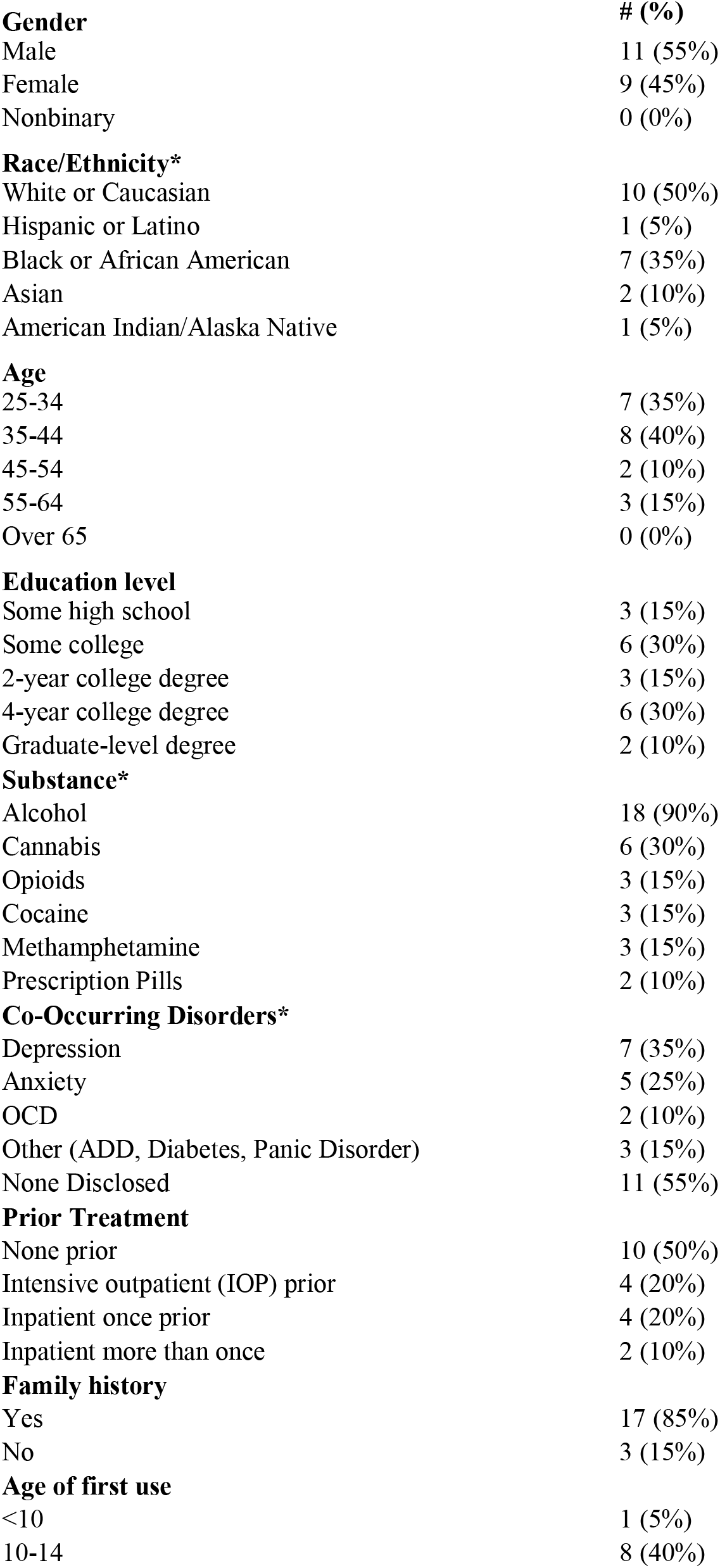
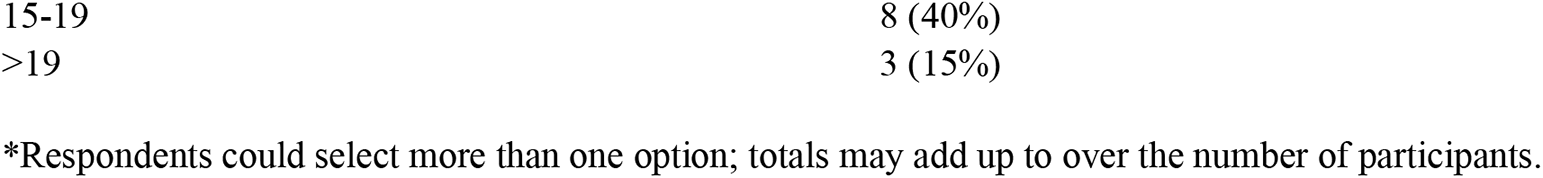
ParticipantDemographics.

| **Gender** | **# (%)** |
| --- | --- |
| Male | 11 (55%) |
| Female | 9 (45%) |
| Nonbinary | 0 (0%) |
| **Race/Ethnicity\*** |  |
| White or Caucasian | 10 (50%) |
| Hispanic or Latino | 1 (5%) |
| Black or African American | 7 (35%) |
| Asian | 2 (10%) |
| American Indian/Alaska Native | 1 (5%) |
| **Age** |  |
| 25-34 | 7 (35%) |
| 35-44 | 8 (40%) |
| 45-54 | 2 (10%) |
| 55-64 | 3 (15%) |
| Over 65 | 0 (0%) |
| **Education level** |  |
| Some high school | 3 (15%) |
| Some college | 6 (30%) |
| 2-year college degree | 3 (15%) |
| 4-year college degree | 6 (30%) |
| Graduate-level degree | 2 (10%) |
| **Substance\*** |  |
| Alcohol | 18 (90%) |
| Cannabis | 6 (30%) |
| Opioids | 3 (15%) |
| Cocaine | 3 (15%) |
| Methamphetamine | 3 (15%) |
| Prescription Pills | 2 (10%) |
| **Co-Occurring Disorders\*** |  |
| Depression | 7 (35%) |
| Anxiety | 5 (25%) |
| OCD | 2 (10%) |
| Other (ADD, Diabetes, Panic Disorder) | 3 (15%) |
| None Disclosed | 11 (55%) |
| **Prior Treatment** |  |
| None prior | 10 (50%) |
| Intensive outpatient (IOP) prior | 4 (20%) |
| Inpatient once prior | 4 (20%) |
| Inpatient more than once | 2 (10%) |
| **Family history** |  |
| Yes | 17 (85%) |
| No | 3 (15%) |
| **Age of first use** |  |
| <10 | 1 (5%) |
| 10-14 | 8 (40%) |
| 15-19 | 8 (40%) |
| >19 | 3 (15%) |

\*Respondents could select more than one option; totals may add up to over the number of participants.

### Findings

#### Initial Reaction to DBS: Unfamiliar and apprehensive, yet interested

Most participants initially expressed unease regarding the physically invasive nature of DBS (i.e., requiring surgery). While two participants had heard of DBS before, none were familiar with how it worked or its current applications. First impressions often described DBS as ‘weird’ and ‘scary’: “wow, it’s crazy, because going deep inside the brain is something that you can’t really play with” (H12) and “Unlike taking an oral pill or taking a shot, it’s invasive. To be honest, that’s a little scary” (H14). Despite that initial unease, when prompted at the conclusion of the interview as to whether or not DBS was something they would ever personally consider, only one participant said no outright: “it reminds me of shock therapy … Oh God. I don’t want something in my head” (H6).

The majority of participants expressed interest, but differed in their perceptions of when DBS would be a reasonable option. Some expressed hesitancy about the exploratory nature of research: “If there’s incentive, yeah I’d do it. But this is research. So it’s dangerous. I’m a little scared” (H20). Many described seeing DBS as a “last resort,” but there was high variation in where people identified that threshold. Some described being open only if they had exhausted all other existing treatment options, while others said they could see themselves reaching last resort desperation with a single relapse: “if I relapse one more time then yeah, I’m all for it” (H16). Others saw themselves as early adopters: “I would definitely raise my hand to say, ‘Hey, let me jump on ship’” (H12).

#### Perspectives on Living with Substance Use Disorder, Treatment, and DBS

Participants described their experiences living with SUDs and in treatment, and how these experiences shaped their perspectives on the prospect of DBS.

#### Living with a Substance Use Disorder (Table 2)

A common theme was that addiction is difficult to overcome and, all too often, fatal. Participants felt a loss of control and at the mercy of their cravings. One participant likened their disorder to a “puppet master” making them do things they could not stop. Participants reported family histories of substance use that often involved recurrent relapse, family trauma, and death.

**Table 2:**
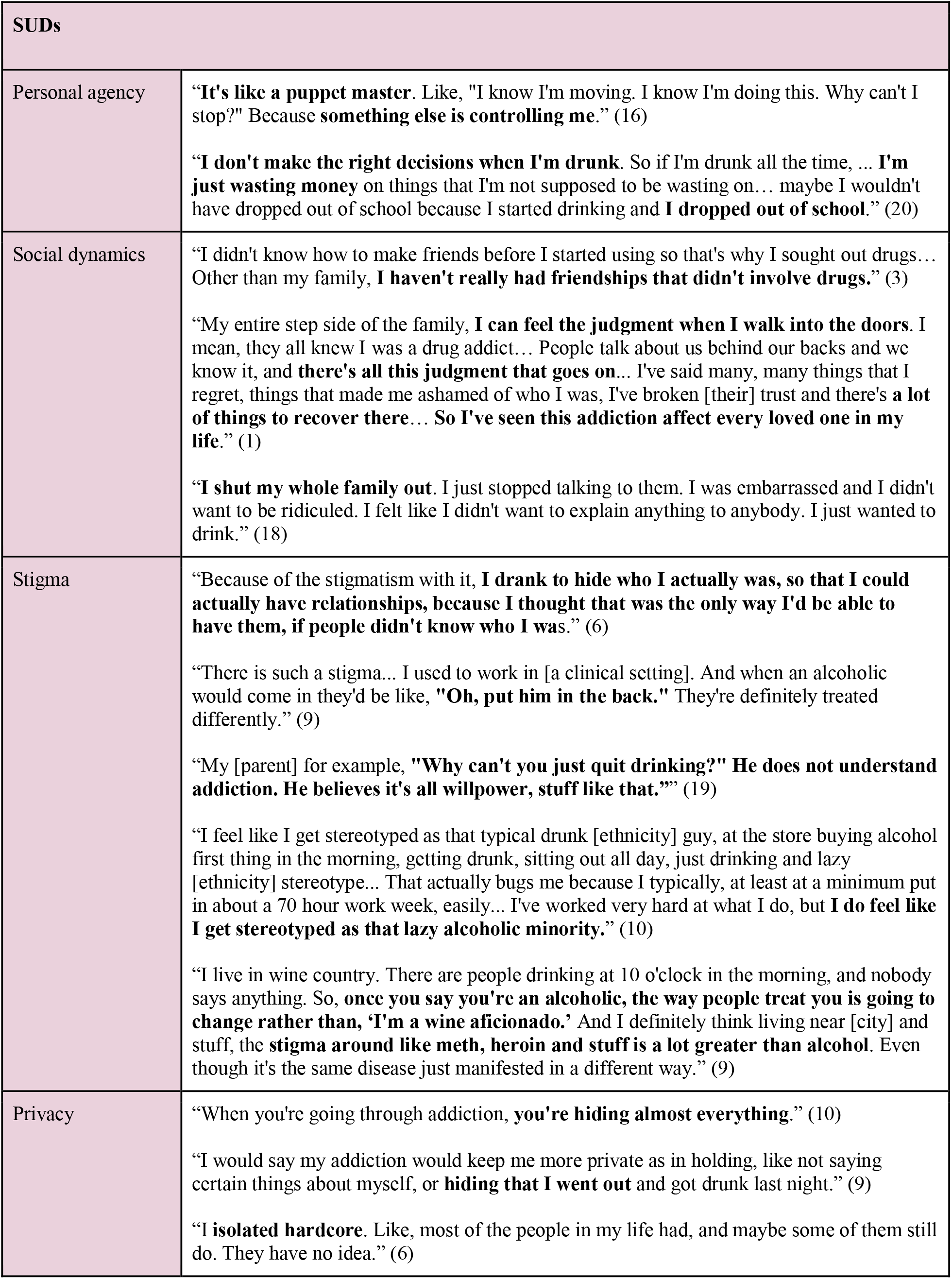
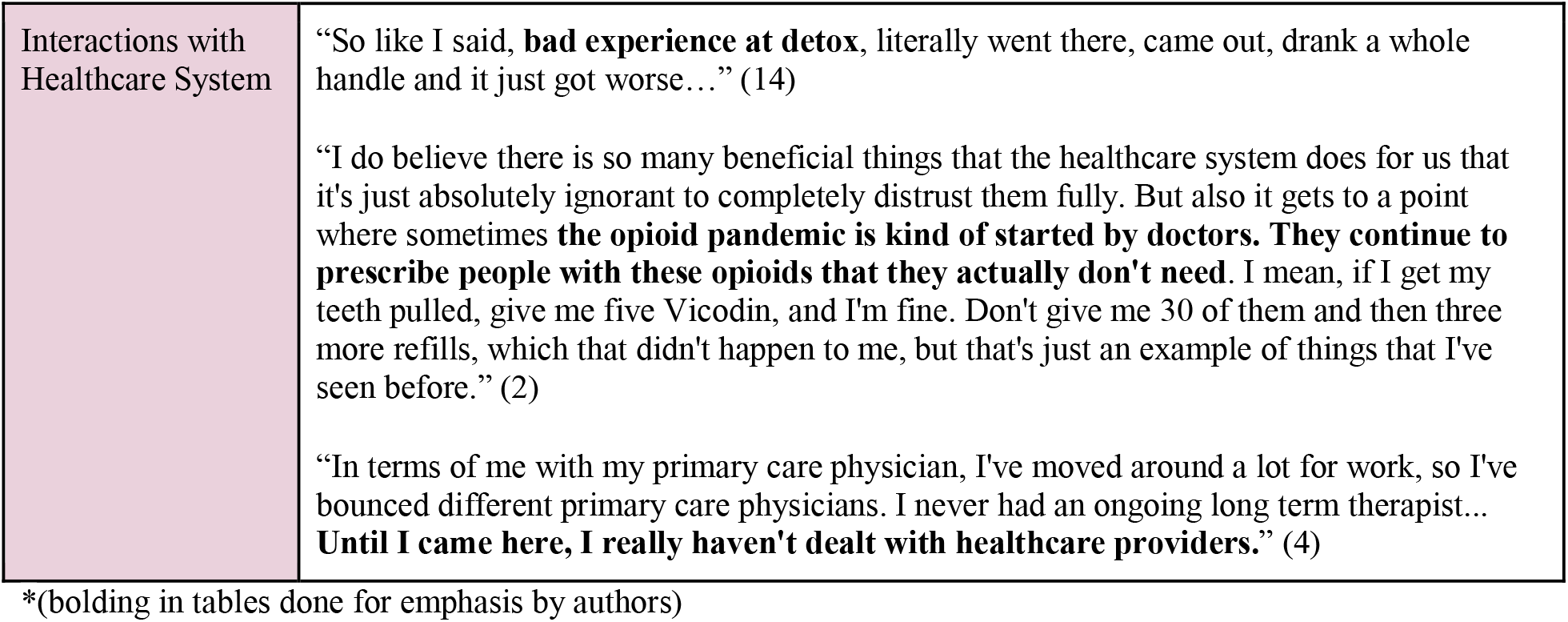
Perspectives on Living with a Substance Use Disorder.

| SUDs |  |
| --- | --- |
| Personal agency | **"It's like a puppet master.** Like, "I know I'm moving. I know I'm doing this. Why can't I stop?" Because **something else is controlling me.**" (16)<br>**"I don't make the right decisions when I'm drunk.** So if I'm drunk all the time, ... **I'm just wasting money** on things that I'm not supposed to be wasting on... maybe I wouldn't have dropped out of school because I started drinking and **I dropped out of school.**" (20) |
| Social dynamics | "I didn't know how to make friends before I started using so that's why I sought out drugs... Other than my family, **I haven't really had friendships that didn't involve drugs.**" (3)<br>"My entire step side of the family, **I can feel the judgment when I walk into the doors.** I mean, they all knew I was a drug addict... People talk about us behind our backs and we know it, and **there's all this judgment that goes on...** I've said many, many things that I regret, things that made me ashamed of who I was, I've broken [their] trust and there's **a lot of things to recover there...** So I've seen this addiction affect every loved one in my life." (1)<br>**"I shut my whole family out.** I just stopped talking to them. I was embarrassed and I didn't want to be ridiculed. I felt like I didn't want to explain anything to anybody. I just wanted to drink." (18) |
| Stigma | "Because of the stigmatism with it, **I drank to hide who I actually was, so that I could actually have relationships, because I thought that was the only way I'd be able to have them, if people didn't know who I was.**" (6)<br>"There is such a stigma... I used to work in [a clinical setting]. And when an alcoholic would come in they'd be like, **"Oh, put him in the back."** They're definitely treated differently." (9)<br>"My [parent] for example, **"Why can't you just quit drinking?" He does not understand addiction. He believes it's all willpower, stuff like that.**" (19)<br>"I feel like I get stereotyped as that typical drunk [ethnicity] guy, at the store buying alcohol first thing in the morning, getting drunk, sitting out all day, just drinking and lazy [ethnicity] stereotype... That actually bugs me because I typically, at least at a minimum put in about a 70 hour work week, easily... I've worked very hard at what I do, but **I do feel like I get stereotyped as that lazy alcoholic minority.**" (10)<br>"I live in wine country. There are people drinking at 10 o'clock in the morning, and nobody says anything. So, **once you say you're an alcoholic, the way people treat you is going to change rather than, 'I'm a wine aficionado.'** And I definitely think living near [city] and stuff, the **stigma around like meth, heroin and stuff is a lot greater than alcohol.** Even though it's the same disease just manifested in a different way." (9) |
| Privacy | "When you're going through addiction, **you're hiding almost everything.**" (10)<br>"I would say my addiction would keep me more private as in holding, like not saying certain things about myself, or **hiding that I went out** and got drunk last night." (9)<br>**"I isolated hardcore.** Like, most of the people in my life had, and maybe some of them still do. They have no idea." (6) |
| Interactions with Healthcare System | “So like I said, **bad experience at detox**, literally went there, came out, drank a whole handle and it just got worse...” (14)<br>“I do believe there is so many beneficial things that the healthcare system does for us that it's just absolutely ignorant to completely distrust them fully. But also it gets to a point where sometimes **the opioid pandemic is kind of started by doctors. They continue to prescribe people with these opioids that they actually don't need.** I mean, if I get my teeth pulled, give me five Vicodin, and I'm fine. Don't give me 30 of them and then three more refills, which that didn't happen to me, but that's just an example of things that I've seen before.” (2)<br>“In terms of me with my primary care physician, I've moved around a lot for work, so I've bounced different primary care physicians. I never had an ongoing long term therapist... **Until I came here, I really haven't dealt with healthcare providers.**” (4) |

\*(bolding in tables done for emphasis by authors)

Participants recounted struggling with shame about ways their SUDs had harmed loved ones. They felt judged by family members and work colleagues. Nearly all had experienced stigma (e.g. SUD understood as a matter of willpower or indicative of moral failure). Most participants struggled to maintain relationships and reported isolating themselves to hide their substance use and avoid stigma. While some participants described how they were before the SUD, many claimed being an “addict” as part of their identity. They shared struggles with self-trust and guilt about manipulating others when the “addict” part of themselves was operative. In addition to the social costs of SUDs, participants described financial and emotional costs, often with negative consequences for personal and work relationships, financial resources, and self-esteem.

#### Perspectives on SUD Treatment (Table 3)

Participants’ sentiments about their treatment and prospect of recovery were often marked by uncertainty, desperation, and determination as they reflected on how much they had lost to their SUD. Some were skeptical of their own ability to recover, given that they had watched peers struggle and relapse. A common concern was uncertainty related to the unpredictability of cravings: “Honestly, I feel like, once I complete treatment, if I were to have a bad enough day, I could potentially say just screw it and go get a drink and snowball back to where I was or even worse” (H19). Nearly all participants viewed SUDs as a long-term disease and recognized their recovery as fragile. As one participant put it, “12 steps is lifelong. It’s forever” (H20). Participants often shared lessons from their 12-step based programs: the need to rely on others, recognition that they could not maintain abstinence alone, and that even with support, they still have to “do the work” to maintain abstinence. Many also mentioned the importance of surrendering to a higher power.

**Table 3:** Perspectives on Substance Use Disorder Treatment.

| Treatment |  |
| --- | --- |
| Personal agency | **“There's some dudes up there, they've been in rehab eight times.** You can see that they really want to try. It's just life hits you hard, bro. Things happen. You get triggered just like you get... This disease is no joke.” (20)<br>“They always tell you, ‘Depend on your higher power.’ Depend on that power that's higher than yourself, because you can't depend on just you. Because if you could depend on just you to get up out of this, you wouldn't be in this situation of addiction. You wouldn't have the relapses. You wouldn't have the cravings or the urges, if you could just depend on yourself. So **apparently we can't just depend on ourselves or rely on ourselves.** We need that help.” (12)<br>“A lot of people that have alcoholism, they probably drink most of their money away. And if they're working to stay on their feet, them getting a job, they better put their sobriety first... But bills don't give a damn about that at the end of the day. You know what I'm saying? **Bills don't give a f\*\*\* about sobriety.**” (20) |
| Social Dynamics | “So I'm **looking forward to mending those relationships sober.** I'm looking forward to that. It's not going to be easy cause I did a lot of wrong stuff and I feel really guilty for a lot of the stuff that I did and I'm going to have to make amends for it and apologize and put myself out there. That's the **hardest part of recovery...** how much you have to put yourself out there so that you can get better, so that you can recover and not drink.” (18) |
| Stigma | “Yeah. I feel like there is. I mean, it's like, **“So-and-So's back in treatment again,”**” (10)<br>“I think **being an inpatient, there's a lot more stigma** than if I would have just said, “I'm going to have to go to therapy for my drinking.” Therapy now is kind of what everybody does.” (9) |
| Privacy | “I know that a **big part of being in recovery is to be honest and open** about our addiction and stuff, but I know **a lot of people that want to be private** about them.” (7)<br>**“My addiction counselor had mentioned, ‘Tell as many people as you can** because the more people that are on board with this, the easier it is because you need your people.’ Because in active use, I isolated so much that I had no one really other than my partner.” (6)<br>**“Just my privacy things.** I'm super concerned, even, to have any mention of this on my medical record, or anything, just because I work in a large hospital, and I know it's easy to get to other people's medical records, and then give them stuff. **I don't want people to think I'm a f\*\*\*ing opioid addict.**” (10) |
| Interactions with Healthcare System | “I have to focus on my money. **It's my life,** as well. What's more important is actually getting my life. I mean, money comes and goes. My life is once. It's once in a lifetime. But it's **always a financial aspect,** especially when you have to take care of a family or when you're responsible for X amount within a household.” (12)<br>**“They give you your antidepressants, you got to come back in six weeks to see if it's working for you and then come back three months later.** I mean, anytime you are doing something or taking something that's supposed to continually help you, but there's still risk of side effects, it's absolutely ignorant to not continue to follow up with your healthcare provider.” (1) |
|  | “I mean, in this fast paced American lifestyle that we live in, everybody's working 40 plus hours a week, you got kids, you got sports activities, schooling and all this stuff that, I mean, sometimes I don't go to the dentist just because I don't have time to go to the dentist. It's the **fear of going into the healthcare system**, but also the **inconvenience of trying to fit it into our busy, crazy daily schedules.**” (1) |

Unprompted, five participants expressed concerns regarding limited treatment options for SUDs. Some had negative experiences with existing treatments (e.g., anti-craving medications with side effects). Financial costs of treatment and the difficulties of finding time in busy schedules were described as burdensome. Participants expressed openness to a variety of methods to achieve recovery, often using the metaphor of “tools in the toolbox” to describe this multi-faceted approach.

While many participants emphasized the importance of understanding and minimizing cravings as a recovery goal, even more participants named building social community and repairing relationships as key recovery aims: “It’s the isolation part of it. It’s crazy because you hear that the opposite of addiction isn’t sobriety. The opposite of addiction is connection. It blows my mind how true that is” (H7). A majority of participants also expressed the desire to gain self-understanding and learn how to better process emotions.

#### Perspectives on Deep Brain Stimulation (Table 4)

Participants expressed concerns regarding DBS risks such as surgical complications. These were often balanced against existing concerns regarding the high risks of relapse and a desire to aggressively avoid that possibility. Many participants saw overcoming cravings as the hardest obstacle to recovery and were drawn to the possibility of DBS to help quiet cravings and understand their patterns: “I am hopeful to get over these cravings eventually to regain control, to find out more about the causes of why I’m like this, kind of like putting myself out there to myself” (H13). Participants who had co-occurring disorders, some of which might also be treatable with DBS, expressed increased interest in DBS if it could simultaneously help them with their depression or OCD.

**Table 4:** Perspectives on Deep Brain Stimulation.

| DBS |  |
| --- | --- |
| Personal agency | “Everybody in the AA community knows that **you can't do it yourself**. And if your higher power, it just happens to be some little magical box that they stuck in your head to help refresh your brain once in a while. I think that would actually be embraced.” (1)<br>“So **now I might have an extra leg up**. I might have an extra tool in my utility belt, but I still got to go out here and do this kung fu... I'm using it, but what if I have a relapse? What if I have a craving or urge? Oh, guess what? I got the device up in me, so it's going to help me with that craving or that urge. And then, from that, I can go back to my book, and I can go back to my steps, or that I can use another tool, my open line of communication with a sponsor, going to a meeting” (12)<br>“You have to put everything on the table and I think that's the hardest part... I'm hoping that I will make amends. I'm willing and **I don't see how that device can do that. You got to kind of, like, make that happen.**” (18)<br>“**I would be afraid that I would get addicted to using it...** one of those videos about what addiction is, it showed the mouse who was pressing the button. He got stimulated, and somehow, I can't remember, it totally overtook everything. So, I just pictured me as a mouse and not drinking, but then not necessarily getting better.” (6)<br>“At this point, I just don't trust my brain to make the right decision at all times. If there's an off chance that I might be able to use it to stimulate it kind of like a drug... **I don't want to end up abusing it.** My goal is to get into recovery and have it last... **I'm just not sure at this point I trust myself quite.**” (15) |
| Social relationships | “I would probably wear one of those as a **badge of honor**, one of those deep brain things just like, “**See, I did everything I could.**”” (9)<br>“If anything, **I'd be happy to show people that I'm willing to do whatever it takes to fix what's going on.**” (11)<br>“My [partner] doesn't have very much experience with it [addiction], so they just don't understand. I know **they just want the madness to stop**. After I finish my phone call with you, I'm going to talk to them about it, and my guess is they would probably applaud me too. Especially if the side effects were minimal, **I think they would push me to do it** [get DBS].” (10)<br>“Having something visible like if she goes and sees her [parent] and her [parent] sees that and she knows what it's for, could just keep making her angry. Just a **reminder that my daughter's a fuck up to her**. I don't know. Or there's something wrong with you. But no, there's not. Whatever.” (7)<br>“**If it can help me, wow, I mean, could it really improve my life, quality of life and my relationships? Can it help me stay out of the depths of self-pity and depression and not finding things in life fun?** I mean, I don't know if it helps with dopamine or anything like that, but can I find things, small things enjoyable or does it always have to be a rush or a high, or something like that?” (15) |
| Stigma | “Would I want something in my head right now, if everything was considered safe and everything? I think I would pick that as an option. I think I would. Yeah. As long as it had a skinny thingy. I got plenty of hair to cover, but yeah... **I don't want to look like Herman** |
|  | **Munster** when I go outside” (14)<br>“And then committing to a device like that, might even take that stigma... **I almost feel like there is more of a stigma on inpatient treatment than there would be with the external device...** Once you attach a medical thing to something, people are like, ‘Well, that’s what my doctor told me to do.’” (10)<br>“There’s **so many people nowadays just so against prescriptions and pills...**’ I can see some stigmas like, ‘You need to go to the extent of getting a device?’ Well, I feel like that’s where it brings light that yes it is serious enough that technology needs to be involved... **I feel like there’d be more stigma against the medication, or more negativity to the medication versus the device,** because obviously the device is a much more serious note.” (19) |
| Privacy | “**If I relapsed,** it’s my choice to figure out what I need to do. **I don’t want anybody like the relapse police running over to my house,** “I know you just relapsed,” or somebody calling me on the phone saying, “You just relapsed and you just had a drink and you wasn’t supposed to.”” (14)<br>“It’s like having a service dog or a service animal and they’re like, ‘Well what is this animal for?’ First of all **you don’t have to disclose that information but if you feel comfortable enough with someone, you will.**” (12)<br>“If I could take care of those cravings, I would’ve preferred not to have to be so open and share so much information about myself... I feel like there’d be, **I’d be able to maintain more privacy with the device.**” (19)<br>“**I wouldn’t want someone to know I have a thingy in my head.** Unless it was because they’re part of my team to make sure I’m safe, to make sure it’s helping me out or whatever... I just always **want to be able to tell my own story.**” (6)<br>“[Law enforcement accessing neural data] That I probably do have an issue with. **I have been manipulated by the justice system several times. I have very little trust in them...** I have several [relatives] doing life sentences... I have seen how they apply pressure to people that have absolutely nothing to do with it... **I’ve never felt a law enforcement officer was there to help me... My trust in law enforcement, the entire system and several officers personally, it’s absolutely zero.** I’d have a huge problem with that, actually.” (11)<br>“So **it’s kind of like a house arrest bracelet,** however you can leave the property and things like that. It just monitors the alcohol intake from your sweat... But I do think that is good because I do need those consequences, otherwise I’m just going to fall right back down the rabbit hole. So **if someone is on probation or some sort of anything involving law enforcement, I do believe they should have access to it [neural data]. However, if it’s just me being a free bird, I don’t think they should be able to.**” (19)<br>“You don’t want that to be used against you in court or something like that. **One of the things people mentioned about George Floyd was his substance abuse and that wasn’t even the major factor.** Despite a person having addictions or whatever it may have been the case, it was wrong, the action. So **you can’t discriminate for that, but people hear something, then they demonize substances.**” (18)<br>“**People feel more comfortable seeing a paramedic more than they would be a police officer,** because when police show up it’s more of an aggressive factor.” (17) |
| Interactions with Healthcare System | “Unfortunately, most alcoholics that are out there are middle-class and poor. **They're not rich.** They don't have a lot of time. Like me, I make okay money, but I got to put in a lot of hours. So like once every two weeks, yeah, I'd agree to it. I'd probably mess around with it, but this is all research, like you said. **How long would the thing take?** How long would it take?” (20)<br>“For me, things like more **logistical things would be of importance.** How long is the study? How often do I have to go see a clinician or a doctor or whatever? Where are they located? Especially, I live in [city], so the **traffic's pretty bad.**” (15)<br>“It's just a thing. Not necessarily good or bad, it's **just part of the treatment.** I don't know. **I already see a therapist regularly,** so... I had to meet with the person that does my psych meds like once a month anyways... I don't think there's any medical treatment that I'm aware of where you don't have to check in with the doctor periodically.” (3)<br>“To me, that's [follow-up clinic visits] **just part of the maintenance of it.** I've only been here a week and they've adjusted my medications three times already... if I personally had it, I could see myself getting irritated without the instant fixing of the cravings I guess. I could see that, but it's not like my [anti-craving medication] NAC, started working immediately.” (19)<br>“I know in the African-American communities, some people would be okay with it. A lot of people wouldn't. **It's just bad connotations with medicine.** A lot of people would point to Tuskegee but it's many incidents. Where the distrust... But **it would be beneficial for people,** all colors, all types because some people feel like they're powerless against drugs and certain substances, especially alcohol.” (18)<br>“**Money talks, bullshit walks, and this is dealing with the brains.** So you're going to have to come out of pocket... it's all about figuring out how to compensate them because putting something in somebody's head and if they have to make out once every two weeks... Most of these people are like me and they got to work. So compensate them.” (20) |

Some participants noted how DBS did not seem drastically different from anti-craving medications, and might even be better: “Honestly the DBS kind of sounds more concrete or reassuring than, because I can have a bad day and just say screw it and not take the pill and then succumb to my craving, whereas I can’t just take that out of my head” (H19). Others found the physical invasiveness potentially off-putting. A couple participants expressed an openness to potentially using the device, but only temporarily, implying that continued reliance on it might be worrisome: “Getting all-natural is definitely the end goal, for sure, but if I need something to kickstart it, all else has failed, so I’m not opposed to it” (11).

Participants viewed the possibility of DBS-related stigma as real, but potentially less concerning than stigma related to other SUD treatment. “I don’t know why they would view me any differently with one of the devices or taking pills. I would say the pros for this device would be, there’s no bottles in my bathroom” (H5). Others expressed that concern regarding the stigma associated with the visibility of the device – scarring, visible wires or battery packs – was lessened due to how visible their disorder had already been to those around them and would be likely counterbalanced by potential benefits.

Nearly all the participants spontaneously mentioned the idea of DBS being another tool in their recovery toolbox. They envisioned DBS as working in tandem with other recovery support systems rather than as a singularly curative intervention. Instead of worrying about the stigma of having a device, for instance, one participant recommended “Being completely upfront with people, saying like, ‘This is simply just a tool as is all of these 12 steps.’ If that’s 12 tools that you have in the 12 steps, this is just my 13th tool that kind of gives me a little bit more help” (H1).

Participants recognized that a neural device that records information could be viewed as a kind of surveillance. Some participants jokingly referenced conspiracy theories about implanted chips and trackers, even as they acknowledged the potential value of allowing health care professionals, and sometimes family members, access to DBS data. Other participants raised concerns about sharing that information, however, even with close family members. Distrust in law enforcement led to most participants not wanting law enforcement officers to ever have access to neural data. Conversely, one participant with experience wearing an alcohol monitoring bracelet had a positive experience with the bracelet being a helpful accountability mechanism, and thought DBS data might serve a similar role.

Some participants were concerned about the financial and time burdens of anticipated DBS programming appointments. Others, however, noted that opioid agonist treatments already often require regular check-ins and the associated burdens of appointments, monitoring, and time off from work. Similarly, participants who worked with therapists were already used to making space in their schedules for regular appointments, and found the regular contact helpful for personal accountability and health maintenance.

#### The importance of patient perspectives for technology development (Table 5)

Reflecting on the interviews themselves, most participants saw them as critical for informing the research process by incorporating perspectives derived from personal experience. Some participants also noted the importance of collecting a wide range of perspectives from people with SUDs, to prevent over-generalization. Other participants framed them as serving an important outreach function of helping inform people with SUDs about the prospect of DBS.

**Table 5:**
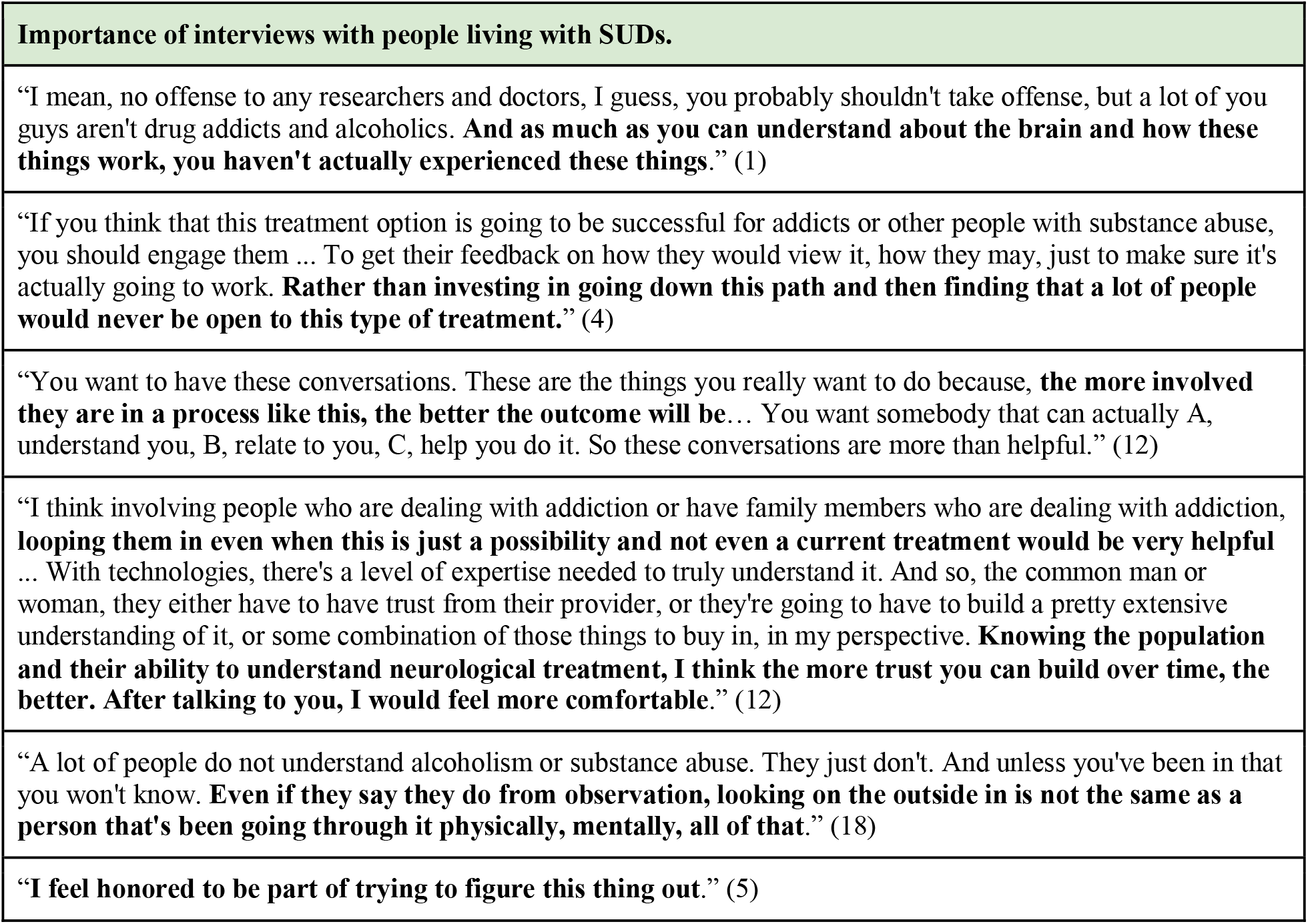
Reflections on the importance of patient perspectives for technology development.

## DISCUSSION

Our results provide evidence that people in treatment for SUDs are open to the possibility of DBS, despite initial apprehensions regarding its physical invasiveness and novelty. This openness often arose from the difficulties and high burdens participants had faced from their SUDs, and their frustrations with access to and effectiveness of existing treatment options. These responses diverged from prior studies of clinicians and researchers that advocate for more restrictive “last resort” criteria (Ali et al. 2016; Lee et al. 2016). Participants viewed SUDs as serious diseases that need better treatment options. In the context of this prior experience, they often considered the risks of DBS well balanced against the potential benefits.

Our findings emphasize t that though DBS may be novel, the considerations it raises surrounding agency, stigma, privacy, and accessibility are not. Participants’ experiences of feeling at the mercy of their cravings led them to be interested in the possibility of DBS helping quiet those urges. Their experiences with the stigma that often surrounds SUD treatment led them to feeling less concerned around the potential stigma of a visually noticeable neural device. Device maintenance appointment burdens were often not viewed as a potential barrier, due to participants’ familiarity with frequent therapy and medical appointments and the 12-step emphasis on needing to “do the work” that recovery requires. DBS was seen as “another tool in the toolbox” that might ease the intensity of cravings, but could never do the complex and expansive work that recovery often requires. Previous work on prospective user acceptability proposes that the degree to which an intervention is considered appropriate depends on the perceived burden, opportunity costs, and ethicality (degree to which it aligns with the user’s value system) (Sekhon et al. 2017). Our findings affirm the relevance of these considerations, and offer additional insight to the way models of acceptability should consider these aspects *relative to* the pre-existing treatment landscape (e.g. opportunity costs of in-patient treatment, 12-step emphasizing “you can’t do it alone”).

These findings also show the importance of consulting those who have the targeted disorder in order to better understand the ethical issues surrounding novel interventions like DBS. Contra Carter et al. (2011), we found that people in SUD treatment, regardless of if for alcohol or opioids, view addiction as “deadly” and available addiction treatments (e.g., anti-craving medications) as either ineffective or causing undesired side effects. Many participants are living with co-occurring disorders that have been proposed as clinical trial exclusions (Ali et al. 2016). Given that many people in our study and the broader SUD population have co-occurring disorders (Han et al. 2017), these exclusions should be reconsidered. The potential reduction of confounding study variables may not be justified if it makes the data inapplicable to the majority of people with severe SUDs (Compton et al. 2007). Indeed, people who are in SUD treatment refractory with frequent relapses may be more likely to live with co-occurring disorders (Najt et al. 2011; Bradizza et al. 2006). There is also reason to expect dual benefit; the most studied DBS target for SUDs (the nucleus accumbens and surrounding white matter) also relieves depressive and anxious symptoms (Sullivan et al. 2021; Widge et al. 2018).

Two additional considerations are highlighted by this study: (1) the importance of increased sensitivity to family dynamics that may complexify caregiver considerations, and (2) the need for increased data protections to prevent further criminalization of SUDs. Participants often named stronger boundary building with loved ones and community rebuilding as goals for recovery. For some, this meant cutting off familial ties and carefully building a new sense of family. Trials of DBS for psychiatric disorders often expect participants to have at least one family member involved with their care and to provide support (Widge & Dougherty 2022). Thus, heightened attention to family dynamics in SUD populations will be needed in clinical trial design.

Secondly, people with SUDs often experience limitations on and threats to privacy due to the degree of criminalization and stigma of SUDs (Kleinman & Morris 2021; AWHONN Position). Limitations on privacy occur, for instance, at the intersection of SUDs and the criminal justice system (e.g., drug monitoring) (Polles et al. 2021; Chang 2020). While much of the discussion of privacy in the context of novel neurotechnology, like DBS, has focused on data security (Bonaci et al. 2014), data ownership (Naufel & Klein 2020), or agency (Schönau et al. 2021), our findings suggest that privacy related to the DBS and the criminal justice system is an underappreciated concern for SUDs specifically. The majority of participants said that law enforcement should never have access to neural data. Both issues require careful consideration should DBS go to clinical trials.

Our study has limitations. Participants were drawn from two clinics within the same treatment system and, notably, all expressed positive experiences with their current treatment program. This experience could have led to more positive appraisals of the potential of DBS, and research more broadly. Further, though we extended recruitment to achieve higher racial diversity, we undersampled many minoritized perspectives, particularly those who hold Asian, Native American, Latino, and Queer identities. Finally, discussion surrounding DBS remained theoretical; no participants had direct experience with the technology, which could have led to stronger framing effects from the video and interview questions. As such, we echo others’ calls for future work to address these perspective gaps (Goering et al. 2022, Wexler & Specker-Sullivan 2021; Shen 2020).

Ultimately, our study shows the importance of understanding novel therapies in the context of the specific features of a disorder, how it is experienced by people who are differently socially positioned, what their treatment options are, and how treatment affects their perspectives on themselves. Addressing the challenges of SUDs will require a multi-pronged approach that makes use of a variety of intervention and support strategies. Participants’ openness to DBS as one “tool in the toolbox” for SUD treatment is notable, but should be considered against the backdrop of substantially unequal access to existing forms of treatment and support, and pressing social problems that contribute to and exacerbate the experience of SUDs. DBS may be able to help some people make significant strides in their recovery, but it cannot address all the broader challenges those with SUDs face in our current social context.

## Data Availability

Data produced in the present study available upon reasonable request to the authors.

## ACKNOWLEDGEMENTS

We thank Ms. Christi Sullivan for filming and editing the DBS briefing video, [anonymized for review] Adam Erdman for helping with interview logistics, and all of our participants who generously offered their time and shared their personal experiences with SUDs and their treatment.

## Notes

### Competing Interest Statement

ASW receives research device donations from Medtronic, which manufacturers deep brain stimulation systems. He receives consulting income from Dandelion Science. He holds multiple granted and pending patents in the general area of deep brain stimulation for psychiatric and substance use disorders, none of which are commercially licensed.

### Funding Statement

This study was funded by NIH P30DA048742 (MJT), R21DA052568 (ASW, MJT), 1RF1MH117800-01 (SG, EK, EV). Additional funding provided by MnDRIVE Brain Conditions Initiative and University of Minnesota Medical Discovery Team on Addiction.

### Author Declarations

Study approved by University of Washington IRB (STUDY00009975).

### Summary of Updates

Manuscript updated to reflect latest paper revisions. Figure 1 revised. Reduced word count for publication.

